# Alternative Qualitative Fit Testing Method for N95 Equivalent Respirators in the Setting of Resource Scarcity at the George Washington University

**DOI:** 10.1101/2020.04.06.20055368

**Authors:** Destie Provenzano, Yuan James Rao, Konstantin Mitic, Sofian N. Obaid, Jeffrey Berger, Sharad Goyal, Murray H. Loew

**Affiliations:** Biomedical Engineering, George Washington University School of Engineering and Applied Science, Washington, DC; Radiation Oncology, George Washington University School of Medicine and Health Sciences, Washington, DC; Department of Anesthesiology and Critical Care Medicine, George Washington University School of Medicine and Health Sciences, Washington, DC

**Keywords:** NIOSH, COVID-19, SARS-CoV-2, Coronavirus, Novel Coronavirus, 3D Printing, N95, Respirator, Mask, Qualitative, Fit Testing, Saccharin

## Abstract

The 2019 Novel Coronavirus (COVID-19) has caused an acute shortage of personal protective equipment (PPE) globally as well as shortage in the ability to test PPE such as respirator fit testing. This limits not only the ability to fit PPE to medical practitioners, but also the ability to rapidly prototype and produce alternative sources of PPE as it is difficult to validate fit. At the George Washington University, we evaluated an easily sourced method of qualitative fit testing using a nebulizer or “atomizer” and a sodium saccharin solution in water. If aerosolized saccharin entered candidate masks due to poor fit or inadequate filtration, then a sweet taste was detected in the mouth of the user. This method was tested against previously fit tested Milwaukee N95 and 3D Printed Reusable N95 Respirator as a positive control. A Chinese sourced KN95, cotton cloth material, and surgical mask were tested as other masks of interest. Sensitivity testing was done with no mask prior to fit test. A sweet taste was detected for both the surgical mask and cotton cloth, demonstrating a lack of seal. However, there was no sweet taste detected for the Milwaukee N95, 3D Printed Reusable N95 Respirator, or Chinese KN95. These results demonstrate this could be a valuable methodology for rapid prototyping, evaluation, and validation of fit in a non-clinical environment for use in creation of PPE. This method should be not be used without confirmation in a formal qualitative or quantitative fit test but can be used to preserve those resources until developers are confident that potential new N95 comparable respirators will pass. We strongly suggest validation of masks and respirators with Occupational Safety and Health Administration (OSHA) approved fit testing prior to use in a clinical environment.

## 1.0 Background

The Occupational Safety and Health Administration (OSHA) defines N95 respirators as tight fitting filtering face piece respirators or filter cartridges that are not resistant to oil, remove at least 95 percent of the most-penetrating particles, and must be fit to the face.^i^ Respirators classified as N95 quality face pieces are therefore self-contained and thought to have a high level of protection from environmental hazards. Because these masks must be air-tight around the face, they must also be fit tested. The 2019 Novel Coronavirus (COVID-19) has caused an unprecedented in the number of practitioners seeking N95 protection and corresponding fit testing. This has resulted in a shortage of not just N95 respirators, but also a shortage in the materials used for fit testing respirators. A survey of 8,200 nurses across the USA reported that only 55% has access to N95 respirators, and only 65% had been fit tested in the last year.^ii^ Commercially available fit tests like those produced by 3M cost $250-350 USD, and are currently on global backorder, and can sometimes take 2 - 3 months to deliver.iii It is notable that a delay of 2-3 months in these resources make the use of this supply chain impractical given that peak resource use due to COVID-19 in the United States is expected on April 15^th^, 2020, which is within several weeks of the writing of this article.^iv^

Considering these shortages, OSHA has instituted temporary guidance to switch from quantitative methods of testing N95 respirators to qualitative methods.v OSH recommends four substances for testing qualitative fit of N95 respirators ^vi^:

- Isoamyl acetate, which smells like bananas;
- Saccharin, which leaves a sweet taste in your mouth;
- Bitrex, which leaves a bitter taste in your mouth; and
- Irritant smoke, which can cause coughing.

Our group at the George Washington University and Hospital determined that there was a need for a qualitative test substitute to continue to rapidly prototype, iterate, and test N95 respirators to compensate for the shortage of PPE. We developed a protocol to mirror OSHA qualitative testing processes and detail it in this report. Our method should be not be used without confirmation in a formal OSHA approved qualitative or quantitative fit test, the method but can be used to preserve those resources until developers are confident that potential new N95 comparable respirators will pass.

## 2.0 Methods

### Current OSHA Recommendations

The United States Occupational Safety and Health Administration defines two official methods for fit testing of N95 equivalent masks: qualitative using four substances that a user can smell or taste, and quantitative that look for negative pressure and aerosol. (OSHA, regulation 1910.134)^vii^ The four qualitative methods as defined above are Isoamyl acetate, which smells like bananas; Saccharin, which leaves a sweet taste in the mouth; Bitrex (denatonium benzoate), which leaves a bitter taste in the mouth; and Irritant smoke, which can cause coughing. OSHA also defines three quantitative methods of generated aerosol, ambient aerosol, and controlled negative pressure.

The fit test involves a sensitivity screening following by a mask fit testing with aerosolized solution as defined by the four parameters above. The sensitivity screening involves placing the subject under a hood without a respirator and aerosolizing one of the four qualitative substances. The subject is asked if they can smell the bananas for Isoamyl acetate, taste the sweet taste for Saccharin, bitter taste for Bitrix, or cough for the smoke. If the fit is positive the subject then puts on the mask and is routinely aerosolized within the hood. While the subject is wearing the mask, they are then instructed to test several conditions including: a) normal breathing b) deep breathing c) turning head side to side d) moving head up and down e) reciting a passage f) jogging in place and e) normal breathing. The test is finished and considered successful if the subject can not taste or smell the substance under any of the aforementioned conditions. If at any point the subject can sense the substance, the test is failed and the fit is judged to be inadequate.

### Aerosolization method

We reviewed the four qualitative methods for the OSHA recommended fit test. Of the four, Saccharin was determined to be low-cost, readily available, and easily detectable. For these reasons Saccharin was selected for candidacy in developing an alternative fit test. We used pellets of sodium saccharin made by NectaSweet, and purchased from a commercial vendor (Amazon, etc.). The tablets contain approximately 30 mg of sodium saccharin each, with a binder of sodium bicarbonate and cellulose. An alternative source may also include “Sweet n Low” packets, which contains approximately 36 mg of sodium saccharin per packet. However, we used NectaSweet tablets because it contained less binder material. NectaSweet tablets were dissolved into a warm neutral water solvent. To form this solution 60 ml water was first warmed to near boiling and then 10 pellets (each containing 30 mg of sodium saccharin) were added to the water and stirred until fully dissolved. The solution was then cooled to room temperature before use. The final solution had a concentration of 5 mg/ml of sodium saccharin.

Commercial OSHA test typically uses a nebulizer to aerosolize particles. We repurposed a nebulizer in the form of an atomizer (a cosmetic spray product available at commercial retailers like Amazon for $10-30 USD) to test fit. A clinical nebulizer used for aerosolizing inhaled medications is also acceptable, if it is available. The subject was instructed to stand within a confined space to mimic the hood. The saccharin solution was loaded into the nebulizer then sprayed first next to the subject, but not directly at the subject, without respirator protection. The subject had to verify they were able to taste the substance before moving to the next step. The subject was instructed to clean their palate of the saccharin with a neutral substance like a glass of water. Next the substance was sprayed around the subject, but not directly at the subject, to verify the subject could not taste the substance through the respirator. Various movements were instructed similar to the commercial fit test such as normal breathing, deep breathing, turning head side to side, moving head up and down, speaking, jogging in place, and normal breathing once again. If the subject could not taste the substance under any of these conditions, the fit of the mask was deemed to be adequate and to pass the test.

### Filtration methods tested

Five filtration and respirator designs were tested using the above saccharin and nebulizer combination. First a subject was instructed to step into a confined space and was sprayed with saccharine solution from the nebulizer. Every subject for every test was able to taste saccharine within fifteen seconds of initial exposure. Next the subject was instructed to drink water to neutralize their palate, then was fit with the respirator. Each mask was tested multiple times with no regard for test order (surgical mask was test before and after the N95 comparable respirators for example to ensure negative result was not false negative).

### The following five methodologies were tested (Figure 1)

1. 3D Printed Reusable N95 Respirator^viii^
2. Kimberly Clark Surgical Mask
3. Powecom KN95 (Sourced from China)
4. Milwaukee N95
5. Hanes Cotton cloth

The 3D Printed Reusable N95 Respirator, KN95, and Milwaukee N95 were all considered N95 or comparable and were first tested for fit and suction prior to test. Subjects placed mask and then performed a qualitative check for any leaks around the surface. The 3D printed Reusable N95 Respirator and Milwaukee N95 respirator had previously passed a qualitative Bitrix fit test at GWUH employee health and functioned as a positive control for the experiment.

**Figure 1:**
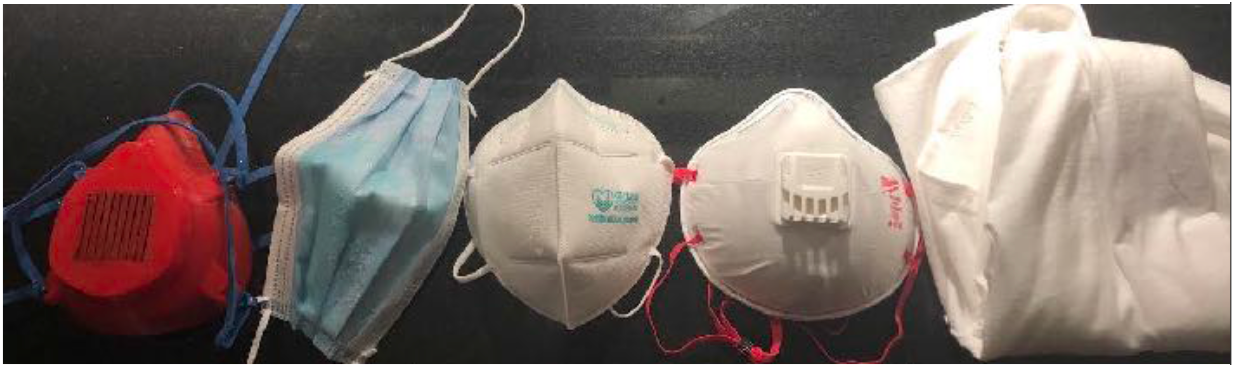
In order from left to right: 3D Printed Reusable N95 Respirator, surgical mask, KN95, N95, cotton cloth.

Cotton cloth and surgical mask were secured prior to test with elastic, however were not considered N95 equivalent and were included as negative controls.

## 3.0 Results

### Qualitative Pre-Assessment of N95 or comparable respirator fit

All three of the N95 or comparable respirators were first visually examined and determined to have no leaks. The 3D printed reusable N95 respirator had the tightest seal, where as the Powecom KN95 appeared to have the least tight seal. Visual inspection of the KN95 revealed perforation along the nose, although this did not seem to ultimately impact the filtration capabilities.

### Qualitative fit test results

Two subjects wore each of five masks two times to have the qualitative saccharin fit test in a confined area.

Both subjects were able to taste the saccharin solution immediately (< 5 seconds of exposure) as a sensitivity test and the test was continued for each respirator. Cotton cloth was fit loosely over subjects’ face and surgical mask was fit according to manufacture instructions to hook elastic around the back of the ears. Neither the cotton cloth nor the surgical mask was able to obtain a qualitative fit (Figure 3):

**Figure 2:**
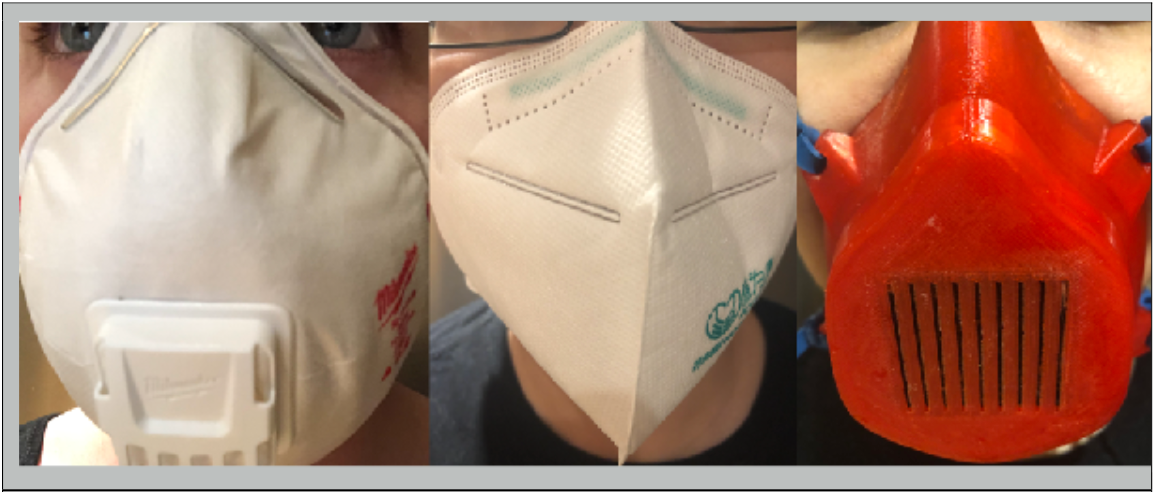
From left to right: Milwaukee N95 Respirator, Powecom KN95, 3D Printed Reusable Respirator. Perforations are visible at the top of middle mask (KN95).

**Figure 3:**
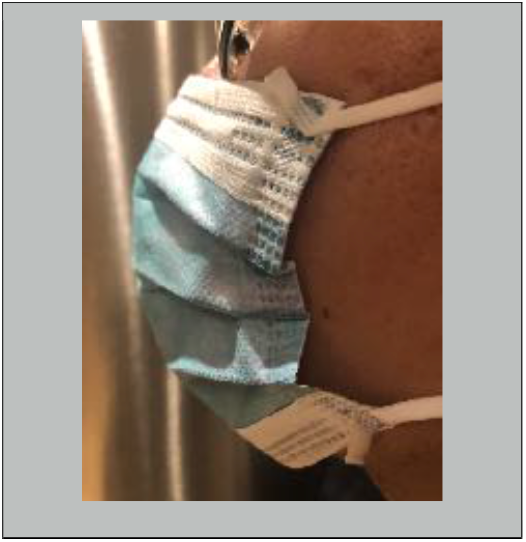
Example lack of suction with Kimberly Clark surgical mask.

Both cloth and surgical mask failed qualitative saccharin solution fit test within 15 seconds of initial exposure and no additional testing was taken. Surgical mask was tested repeatedly to ensure this was not a false result. On average, sweet taste was detected within 45 seconds with a surgical mask or cotton cloth.

All three of the remaining respirators resulted in no detectable taste of the saccharin solution by the wearer. The KN95 mask despite initial pre-assessment qualitative appearances, withstood multiple tests and movements and did not allow any particulates through for taste. It should be noted that the N95 as well as the 3D printed reusable N95 respirator had previously passed a qualitative Bitrix fit test at GWUH employee health (Results not shown).

The 3D Printed Reusable Respirator was tested with MERV 16 filter wrapped in MERV 13, which previously passed the Bitrix fit test. Subject reported no ability to taste saccharine while the respirator and filter were in place. Once the cartridge was removed, the subject was able to rapidly discern the saccharin.

## 4.0 Discussion

The process described above demonstrated that it is possible to emulate a clinical qualitative fit test using easily available materials. The sensitivity test was detectable for all tests of all masks and all subjects, suggesting the atomizer and saccharin solution functioned comparable to a nebulizer for this purpose similar to clinically used Saccharin or Bitrix in qualitative fit tests. The N95 respirator has been approved by OSHA and US NIOSH, functioning as high efficiency particulate filter against coronavirus, and a positive control for this study. (OSHA 29CFR1910.134) The 3D Printed Reusable N95 Respirator had been previously tested in employee health, so it also served as a positive control for this purpose.

Most interesting was the KN95. The KN95 mask is outsourced from China and was approved for use in the USA by the CDC in the setting of resource shortage caused by COVID-19.^ix^ (CDC, GB/T 18664— 2002). This test passed the qualitative fit test described in this report, but has not yet been evaluated with a OSHA certified fit test at GWUH.

The cloth mask provided the least protection against detection of saccharin aerosols and can not be expected to be a tight sealing mask. This result was expected, but is of particular interest due to widespread creation of cloth masks in the community. Additionally the Kimberly Clark surgical mask also did not pass the seal test and also demonstrates that it is not adequate protection to aerosol particulates. However, both masks resulted in some delay in the detection of saccharin aerosol compared to no mask at all. Although it cannot be assumed that either cotton cloth nor surgical mask would provide complete protection against aerosols like the 2019 Novel Coronavirus, they are likely better than not wearing any mask at all.

These results suggest similar tests could be used to rapidly prototype and fit N95 comparable respirators outside of a clinical environment due to the shortage of PPE, personnel, and access to clinical grade fit testing. It should be emphasized that after passing the protocol described in this study, clinical fit testing should still be performed and passed before N95 equivalent masks or respirators are put into use with COVID 19 positive patients. However, this study does allow for rapid testing and cycling of designs and may speed development of PPE during the COVID-19 pandemic in the setting of resource limitations.

## 5.0 Conclusion

Aerosols created using a nebulizer and an easily sourced saccharin solution were able to be used to test the fit of several different N95 equivalent and non-equivalent masks and respirators. This could provide a valuable non-clinical grade evaluation of N95 respirators to rapidly develop PPE during the COVID-19 pandemic. We strongly suggest validation of masks and respirators with OSHA approved fit testing prior to use in a clinical environment.

## Data Availability

All data used in this study is posted and freely available to the public. Any additional files are available upon request from the authors.

## Notes

Conflict of interest: None

### Competing Interest Statement

The authors have declared no competing interest.

## References

i https://www.osha.gov/video/respiratory_protection/resptypes_transcript.html

ii National Nurses Association: “NNU COVID-19 Survey Results.” Survey. March 31, 2020.

iii https://www.envirosafetyproducts.com/3m-bitrex-qualitative-fit-test-kit.html

iv https://covid19.healthdata.org/projections

v https://www.osha.gov/news/newsreleases/national/03142020

vi https://www.osha.gov/video/respiratory_protection/fittesting_transcript.html

vii https://www.osha.gov/video/respiratory_protection/fittesting_transcript.html

viii Provenzano, D.; Rao, Y.J.; Mitic, K.; Obaid, S.N.; Pierce, D.; Huckenpahler, J.; Berger, J.; Goyal, S.; Loew, M.H. Rapid Prototyping of Reusable 3D-Printed N95 Equivalent Respirators at the George Washington University. Preprints 2020, 2020030444 (doi: 10.20944/preprints202003.0444.v1).

ix https://www.cdc.gov/coronavirus/2019-ncov/hcp/respirators-strategy/crisis-alternate-strategies.html

